# Susceptible supply limits the role of climate in the COVID-19 pandemic

**DOI:** 10.1101/2020.04.03.20052787

**Authors:** Rachel E. Baker, Wenchang Yang, Gabriel A. Vecchi, C. Jessica E. Metcalf, Bryan T. Grenfell

## Abstract

Preliminary evidence suggests that climate may modulate the transmission of SARS-CoV-2. Yet it remains unclear whether seasonal and geographic variations in climate can substantially alter the pandemic trajectory, given high susceptibility is a core driver. Here, we use a climate-dependent epidemic model to simulate the SARS-CoV-2 pandemic probing different scenarios of climate-dependence based on known coronavirus biology. We find that while variations in humidity may be important for endemic infections, during the pandemic stage of an emerging pathogen such as SARS-CoV-2 climate may drive only modest changes to pandemic size and duration. Our results suggest that, in the absence of effective control measures, significant cases in the coming months are likely to occur in more humid (warmer) climates, irrespective of the climate-dependence of transmission and that summer temperatures will not substantially limit pandemic growth.

## Main text

The SARS-CoV-2 pandemic represents a generational public health concern. Sustained local transmission is present in multiple countries, in all continents, and the implications in terms of morbidity and mortality are expected to be severe [1, 2]. The role of seasonal and geographic climate variations in modulating the transmission of the virus has received increasing attention. Studies using a regression framework have found a role for temperature, relative humidity and specific humidity in the transmission of SARS-CoV-2 [3, 4, 5, 6, 7], suggesting that cold, dry conditions increase the transmission of the virus. However, with limited data on the current epidemic, these early-stage results are inevitably inconclusive. Furthermore, the relative importance of climate drivers when compared to high population susceptibility during the pandemic stage of an emerging infection such as SARS-CoV-2, has not been fully characterized.

Climate affects the transmission of several directly-transmitted pathogens [8]. Specific humidity has been shown to be important for influenza transmission in both laboratory settings [9, 10, 11], as well as in population-level studies [12]. Respiratory syncytial virus (RSV), a childhood pathogen, has also been found to be dependent on specific humidity [13] and exhibits latitudinal correlations with climate [14]. For both influenza and RSV, low specific humidity increases transmission and epidemics tend to peak in the wintertime in northern latitudes. However, other directly-transmitted infections exhibit different patterns [15], with enteroviruses, for instance, often peaking in the summer months [16].

Prior work on climate and directly-transmitted diseases has typically considered endemic infections, such as seasonal influenza or RSV. Emerging pathogens, in contrast, have distinct dynamics driven by high population susceptibility. A key question is the extent to which seasonal and geographic climate variations are relevant in the pandemic phase of an emerging infection. Here we build on known features of coronaviruses and other directly-transmitted infections to probe this question. Although we do not yet know the climate sensitivity of SARS-CoV-2 transmission directly, data exists on four other coronaviruses that currently circulate in human populations. Two of these coronaviruses, HCoV-HKU1 and HCoV-OC43, are of the same betacoronavirus genus as SARS-CoV-2 [17].

We use data on HCoV-HKU1 and HCoV-OC43 from US census regions to understand the potential climate dependence of betacoronavirus transmission [18]. We fit a Susceptible-Infected-Recovered-Susceptible (SIRS) model to case data of HCoV-HKU1 and HCoV-OC43 where the fitted parameters include the climate dependence of transmission and the length of immunity following infection. All other parameters are fixed, based on values from Kissler et al 2020 [17]. Motivated by the climate-dependence of influenza and RSV, we posit that transmission depends on specific humidity: we use population-weighted average climatology of specific humidity over 2014-2020 taken from the ERA5 reanalysis dataset [19], with population data from [20].

After fitting the model parameters, we run simulations of the SARS-CoV-2 pandemic under three scenarios. In the first scenario we assume SARS-CoV-2 has the same sensitivity to climate as influenza, based on a prior model from laboratory studies [9, 12]. In the second and third scenarios, we assume SARS-CoV-2 has the same climate dependence and length of immunity as HCoV-OC43 and HCoV-HKU1 respectively. While we assume the climate dependence is the same as these three infections, our simulations use R0 based on current estimates of SARS-CoV-2 [17].

## Results

We first look at the seasonality of the endemic betacoronaviruses. Fig.1 shows average monthly specific humidity and normalized cases (between 0 and 1) for OC43 and HKU1 in the US. Cases of both diseases increase as specific humidity declines. For comparison, we show the relationship between specific humidity and SARS-CoV-2 in Wuhan, China during the first 9 weeks of 2020. In contrast to the endemic coronaviruses, cases of COVID-19 shown an *increase* with humidity. As explored below, this does not mean that high humidity increases transmission, but that pandemics driven by novel pathogens may exhibit distinct patterns compared to endemic diseases [21]. We assume that to some extent transmission will decline with specific humidity, however, the extent of the decline is yet to be determined. We characterize the link between specific humidity and the transmission of SARS-CoV-2 using plausible estimates derived from the two endemic betacoronaviruses as well as influenza. Fig.2 shows different potential functional forms for the climate-transmission relationship. Changes to specific humidity modulate *R*_0_ between a maximum wintertime value and a hypothesized lower bound, taken from prior studies [17]. In the extreme cases, transmission (*R*_0_) either rapidly declines as specific humidity increases or has no relationship with specific humidity. The highlighted influenza relationship is based on laboratory studies using the guinea pig animal model [9, 11, 10], and later used to predict influenza epidemics in human populations [12]. In this case *R*_0_ values correspond to SARS-CoV-2 estimates.

**Figure 1:**
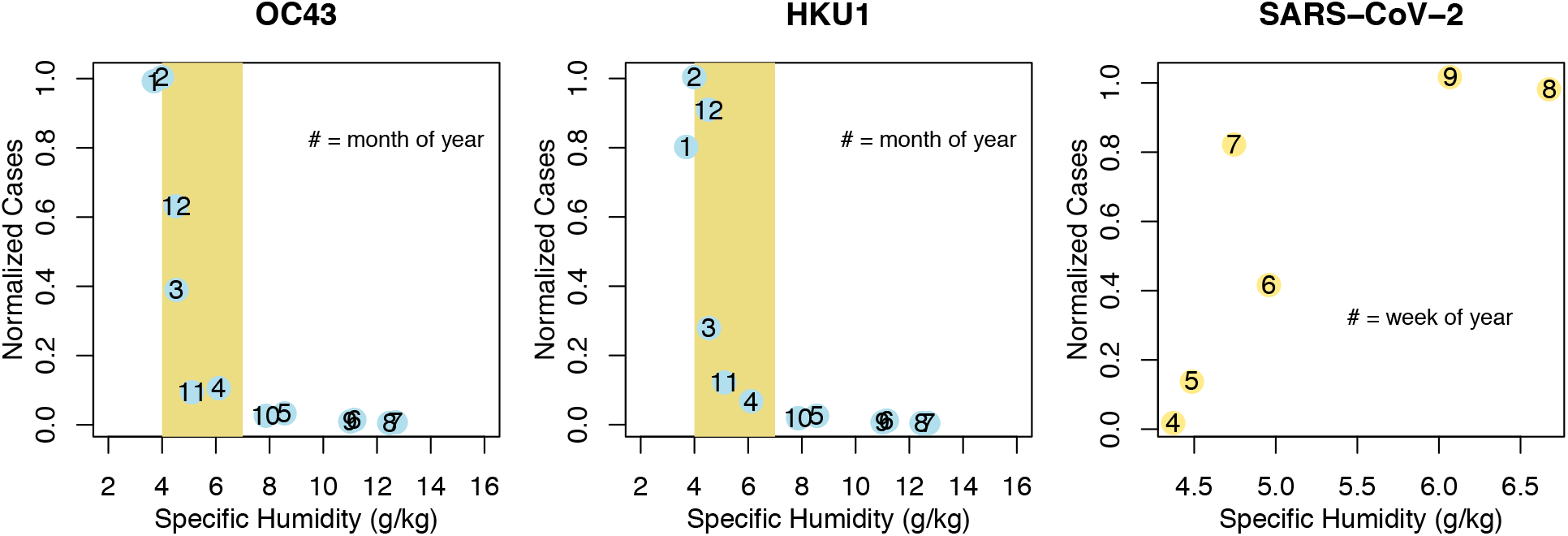
Seasonality of coronaviruses. Average monthly specific humidity plotted against normalized (between 0 and 1) cases of OC43 and HKU1 averaged across the US from 2014-2020, as well as normalized cases of SARS-CoV-2 in Wuhan from weeks 4-9 of 2020. The yellow shaded in area in the first two plots corresponds to the width of the x-axis in the final plot, for comparison.

**Figure 2:**
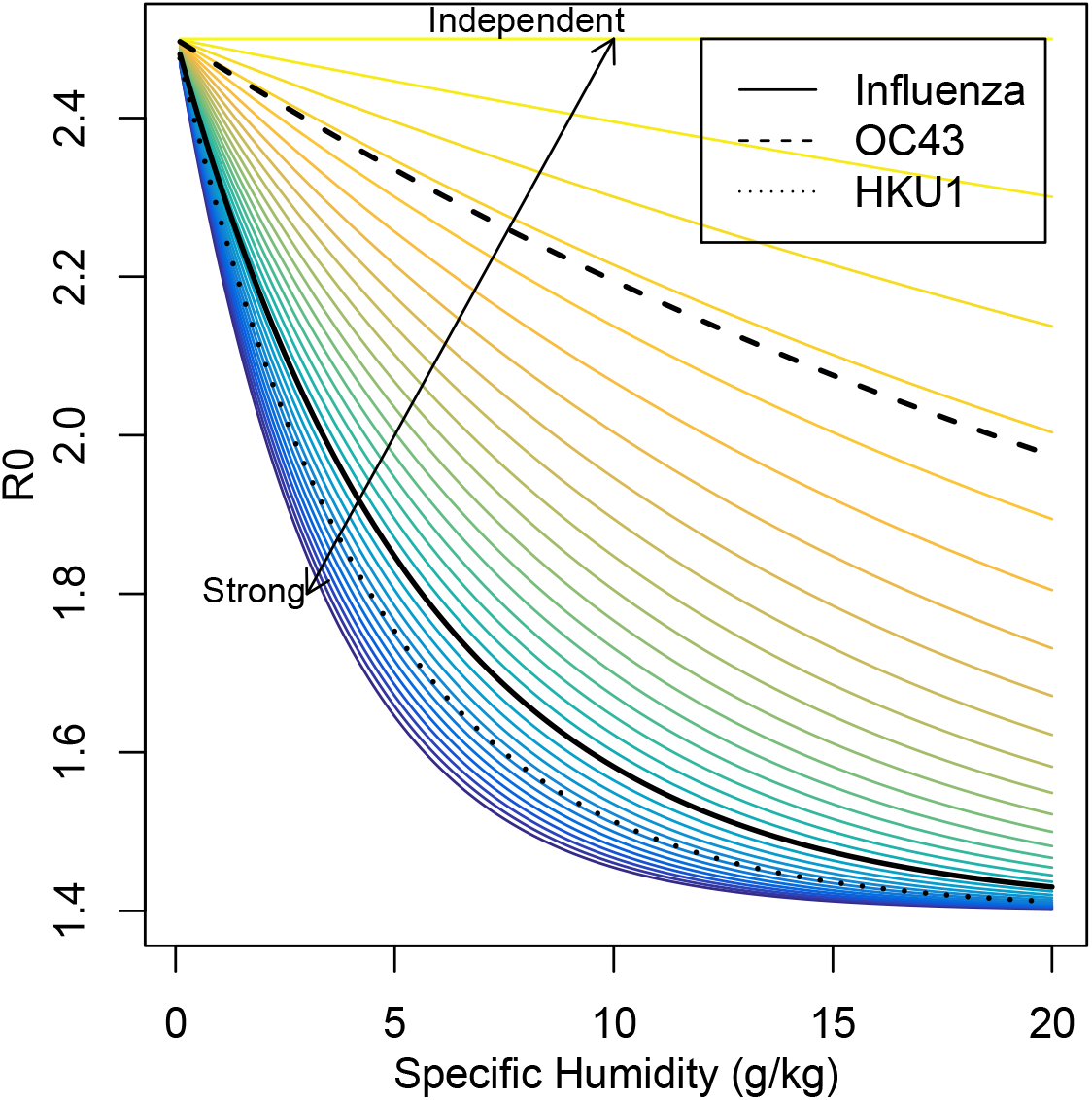
Specific humidity and transmission. The colored lines represent different hypotheses for the relationship between climate and transmission for SARS-CoV-2. The relationship between climate and influenza transmission, OC43 transmission and HKU1 transmission are shown with black lines.

The other two scenarios in Fig.2 correspond to the relationship between climate and OC43 and HKU1 transmission. We evaluate the functional form of this relationship by fitting our climate-driven SIRS model to US case data for the two infections (Fig.S1 and S2). Our results (Fig. 2) suggest a somewhat wide range of climate dependency for the two coronaviruses, with HKU1 having a much steeper response to specific humidity when compared to OC43. Strong seasonal forcing has been linked to biennial outbreaks, as observed for HKU1, in other respiratory pathogens [13] and implies dynamics driven by herd immunity; however this inference may be complicated by cross-protection from other circulating strains [17]. While there is some uncertainty in our estimates, simulating a pandemic outbreak using a range of climate-transmission dependencies allows us to explore a wide plausible range of potential climate effects.

We simulate a pandemic invasion using the climatology of nine exemplar cities each with a very distinct mean and seasonal cycle of specific humidity (3a,b). We stress that these simulations explore only the interaction of the epidemic (SIRS) model clockwork and seasonality in transmission; they do not address complexities of demography, control and other environmental factors. In Fig.3c-e we show the evolution of the simulated pandemic, holding population constant, for northern hemisphere, southern hemisphere and tropical locations. The model assumes the outbreak starts at the same time and no control measures are in place, revealing only the effect of climate on pandemic size and duration. For the northern hemisphere locations, we do not see any substantial difference in pandemic size across all three scenarios, despite very different climates in New York, London and Delhi. In the influenza and HKU1 scenarios, tropical locations experience a more sustained, lower intensity pandemic than the northern hemisphere. These scenarios represent a stronger dependence on climate than OC43, such that the lack of really dry conditions (low specific humidity) in tropical regions means these locations do not experience the high transmission rates of New York, London and Delhi. However, the outbreak in the tropical cities remains significant, and factors we do not explore here, such as population density, could further exacerbate the size of the epidemic.

**Figure 3:**
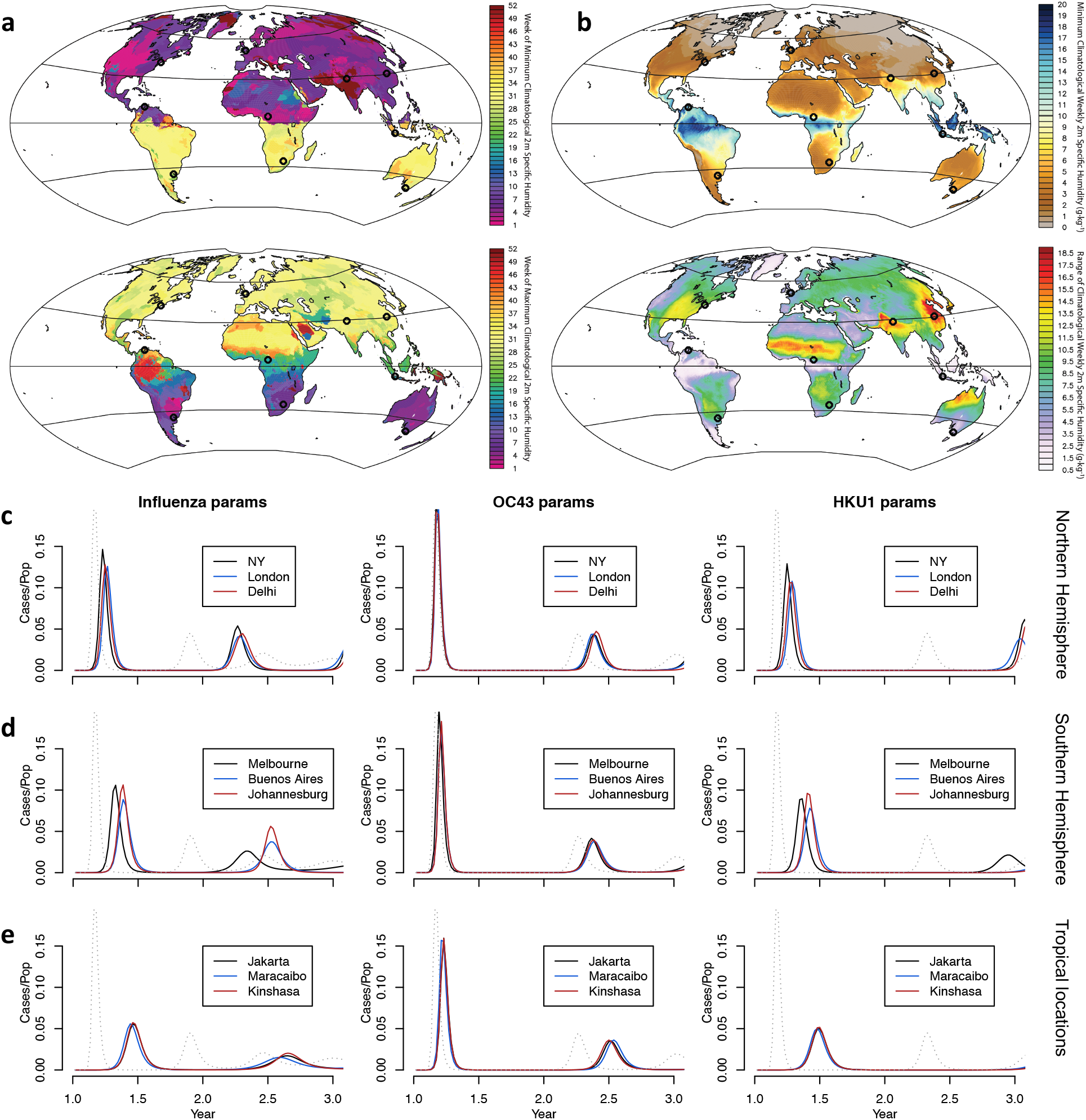
Climatology and model results for nine locations. The timing of minimum and maximum specific humidity (a) and minimum/ humidity range (b) is shown for the globe. Locations where we run simulations of the pandemic are shown with circles. Simulated pandemics are shown for cities in c) the northern hemisphere, d) the southern hemisphere and e) tropical locations. The dotted line represents a pandemic with no climate dependence.

We also simulate the pandemic in a range of southern hemisphere locations (Fig.3d). We do not see a delay in the peak of southern hemisphere locations, despite distinct seasonality when compared to the northern hemisphere. For the OC43 scenario, pandemics are temporally aligned across all locations and of similar magnitude. A stronger climate response for influenza and HKU1 parameters leads to slight regional differences. It is worth noting that our different scenarios also reflect a range of immunity lengths. The size of the pandemic peak is not affected by changes in immunity length, but the timing of latter stage outbreaks is partially dictated by this parameter. The differential timing of secondary peaks in the influenza and HKU1 scenarios, which have a similar climate-dependence, is driven by this variability.

During the pandemic stage of an emerging pathogen the lack of population immunity, i.e. high susceptibility, is a crucial driver. To illustrate this in the general case, we run our simulation model for different climates (represented by the seasonal range of humidity values a location experiences) and different levels of population susceptibility. Fig.4 shows the results in terms of the size of the summertime pandemic peak. While humidity range does modulate pandemic size, population susceptibility exhibits a much steeper gradient. For novel pathogens, such as SARS-CoV-2, the proportion of the population susceptible to infection may be close to 1. To illustrate the potential longer term behaviour of the pandemic, we plot a typical pandemic trajectory on the SI phase plane (Fig.5). The initial pandemic trajectory (red) is relatively independent of seasonal forcing. This then gives way to the endemic attractor (blue) which oscillates around the equilibrium of the unforced model (green). These longer term dynamics show a much stronger signature of seasonal forcing than the pandemic phase. Note that we ignore the effects of control measures and potential coronaviral competition in the simulations.

**Figure 4:**
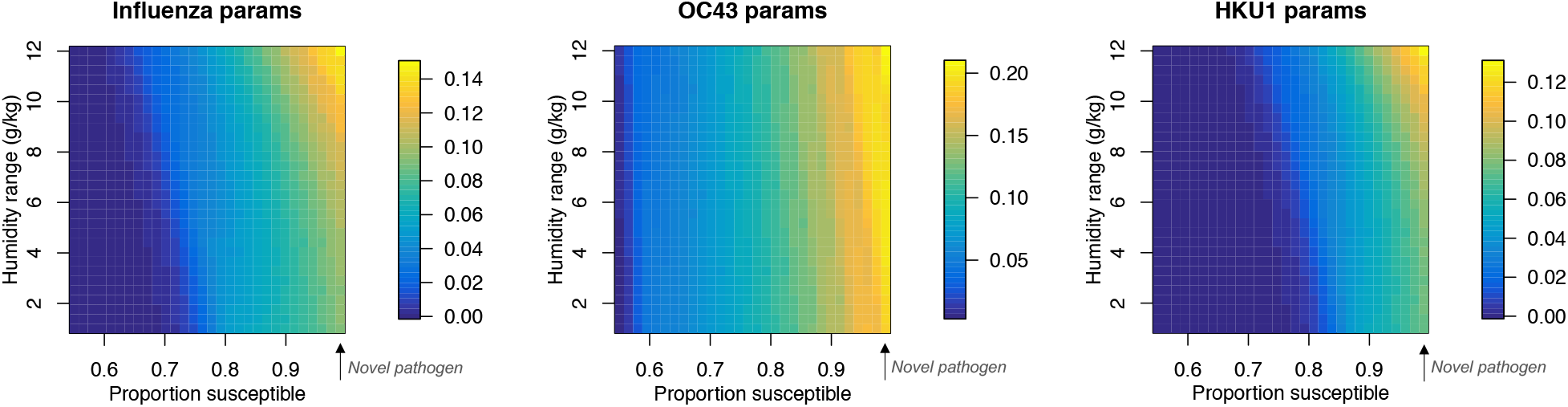
Pandemic peak size depends on proportion susceptible. For the three scenarios (influenza, OC43 and HKU1) the surface plot shows the dependence of maximum pandemic incidence/capita on seasonal range of humidity and proportion of population susceptible.

**Figure 5:**
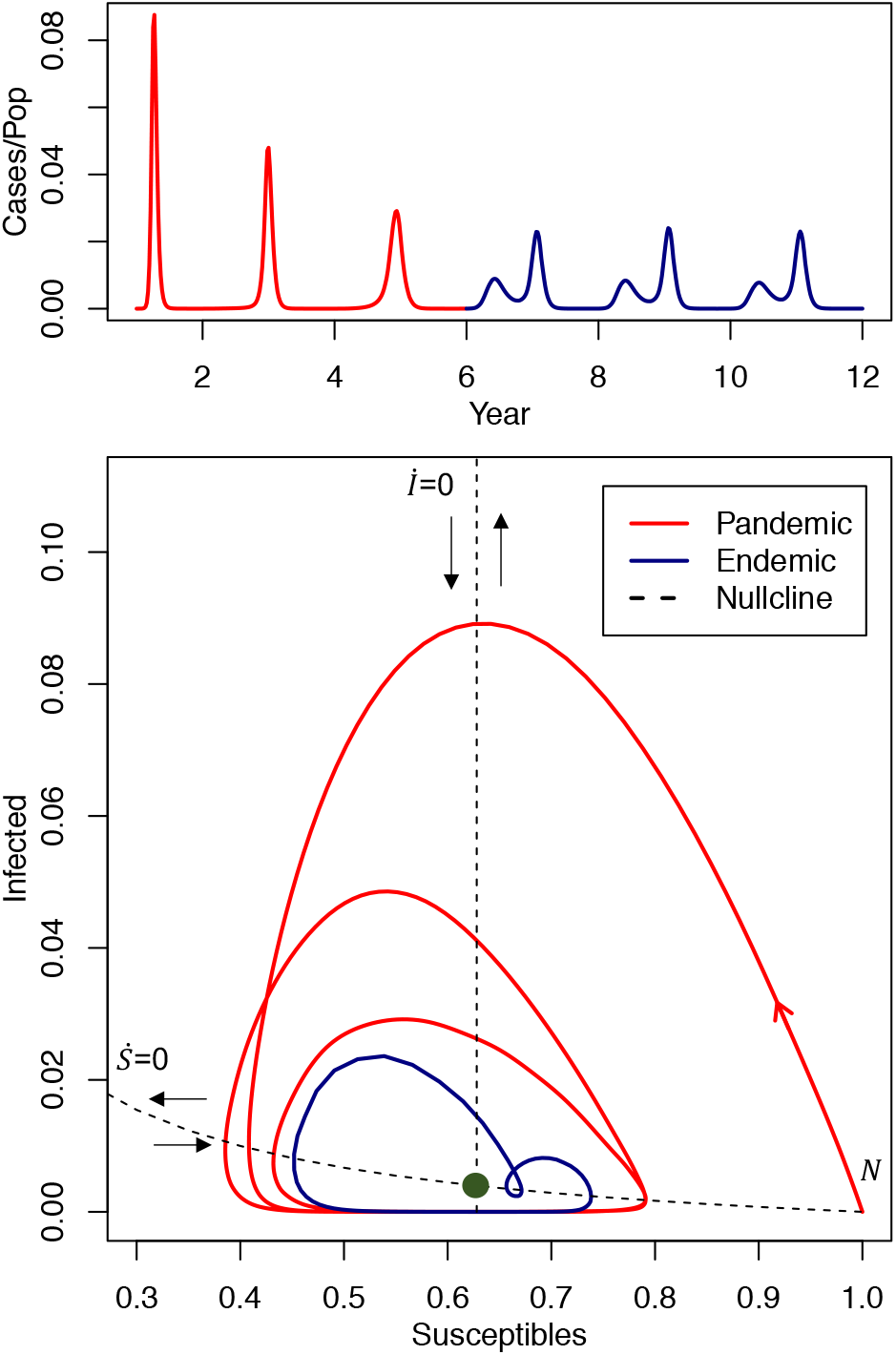
SI phase plane for pandemic and epidemic trajectories. Lower plot: phase plane with trajectory based on Wuhan climatology, using influenza parameters. The two nullclines are from the unforced SIRS using mean *R*_0_. The green circle represents the equilibrium of the unforced model. Upper plot: equivalent epidemic time series.

## Discussion

There are several caveats to interpreting these results. Primarily, our simulations do not address control measures, such as social-distancing, that are being implemented in many countries at present. Our model also does not account for potential cross-protection from other coronavirus infections [17]. Finally, results from influenza and RSV suggest that high precipitation may play a role in driving transmission [13, 22], particularly in tropical locations. Due to limited data on betacoronaviruses from tropical locations, we have not been able to confirm whether a rainfall signal exists.

Our results suggest that while climate may play a role in modulating detailed aspects of the size and timescales of a pandemic outbreak within a particular location, population immunity is a much more fundamental driver. Although our HKU1 scenarios presents a modest role for climate in terms of shifting the timing and intensity of the pandemic, a scenario with OC43 parameters is equally likely. In terms of the SARS-CoV-2 pandemic, our results imply that both tropical and temperate locations should prepare for severe outbreaks of the disease and that summertime temperatures will not effectively limit the spread the infection. However, this does not mean the climate is not important in the longer term. Endemic cycles of the disease will likely be tied to climate factors and seasonal peaks may vary with latitude (Fig.S3). A more detailed understanding of climate drivers as well as immunity length will be crucial for understanding secondary outbreaks. Furthermore, weather and near-term climate forecasts could be helpful for predicting such outbreaks after the initial pandemic phase has passed.

## Methods

### Data

Data on endemic coronaviruses come from The National Respiratory and Enteric Virus Surveillance System (NREVSS) [18] and are available on application after signing a data-use agreement with the CDC. Following [17], we multiply percent positive test numbers by the proportion of weekly ILI visits obtained from the US Outpatient Influenza-like Illness Surveillance Network (ILINet).

Climate data for the US regions come from ERA5 [19] and population-weighted averages of specific humidity over the census region area are constructed using 2015 population data from CIESIN [20]. We calculate the average climatology for 2014-2020 to the run the model and do not consider year-to-year climate variations. For simulating global pandemics, we calculate a 30 year climatology using specific humidity from NASA’s Modern-Era Retrospective analysis for Research and Applications (MERRA) dataset.

### Model

The climate-driven SIRS model is based on [12]:

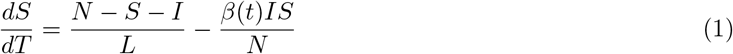

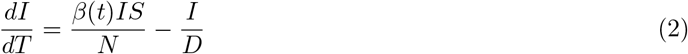

where *S* is the susceptible population, *I* is the number of infectious individuals and *N* is the population. *L* represents the duration of immunity and *D* is the mean infectious period. *β*(*t*) is the contact rate at time *t* and is related to the basic reproductive number by *R*_0_(*t*) = *β*(*t*)*D. R*_0_ is related to specific humidity *q*(*t*) using the equation:

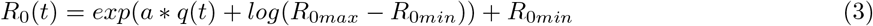

where *a* is the climate dependence parameter and *R*_0*max*_, *R*_0*min*_ are the maximum and minimum reproductive numbers respectively. We assume a flat reporting rate which is used to scale model output to cases.

For OC43 and HKU1 we find *a* and *L* by fitting the model to data from census regions in the US. We take the average fit across census regions to represent the OC43 and HKU1 values of *a* and *L*. In general, parameter values for each disease show agreement across census regions. The western region is a slight outlier for both diseases. As this region pools data from both Alaska and Hawaii (as well as western US states) it represents a large range of climatologies and this likely makes the true climate effect harder to disentangle.

We fit over a range of climate dependencies reflecting no climate dependence to approximately double the known climate dependence on influenza [0,300]. The immunity length of SARS-CoV-2 is yet to be determined, however, some early stage research found that rhesus macaques could not be reinfected 30 days post-infection [23]. Studies using another endemic coronavirus, NL63, found that neutralizing antibody titers declined to zero 52 weeks after first infection [24]. Using the US betacoronavirus data, another modelling effort found an immunity length of 40 weeks [17]. Here, we assume 20 weeks in the lower bound on immunity length, and allow the model to fit a maximum immunity length of two years. Our fitted immunity lengths, *L*, are 62.5 and 66.25 weeks for OC43 and HKU1 respectively. Our fitted values for *a* are -32.5 and -227.5 for OC43 and HKU1 respectively. Visualization of the fitting process is shown in Fig. S1 and S2.

## Data Availability

US coronavirus data is available from the The National Respiratory and Enteric Virus Surveillance System (NREVSS) on request. Climate data is publicly available from ERA5 and NASA.

https://www.ecmwf.int/en/forecasts/datasets/reanalysis-datasets/era5

https://gmao.gsfc.nasa.gov/reanalysis/MERRA/.

## 1 Supplemental Figures

**Figure 1:**
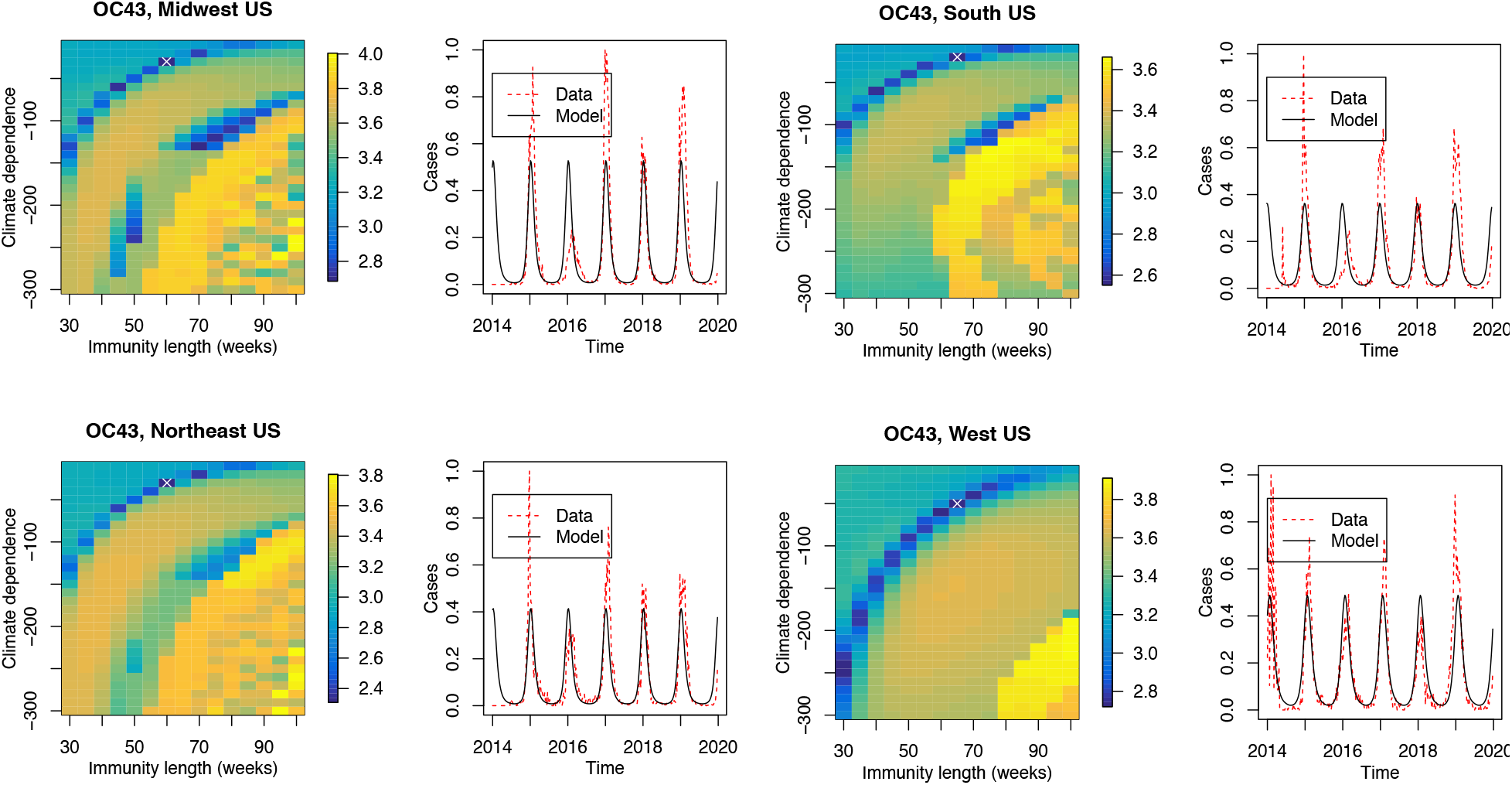
Model fit for OC43. Two parameters are fitted (climate dependence and immunity length) using case data for OC43 in four US census regions. Our model fits only the average seasonality of OC43 cases and does not take into account year-to-year variation. The surface plot shows the log of the sum of absolute error. Darker regions represent a better fit of our model and the best fit parameters are shown with a white cross.

**Figure 2:**
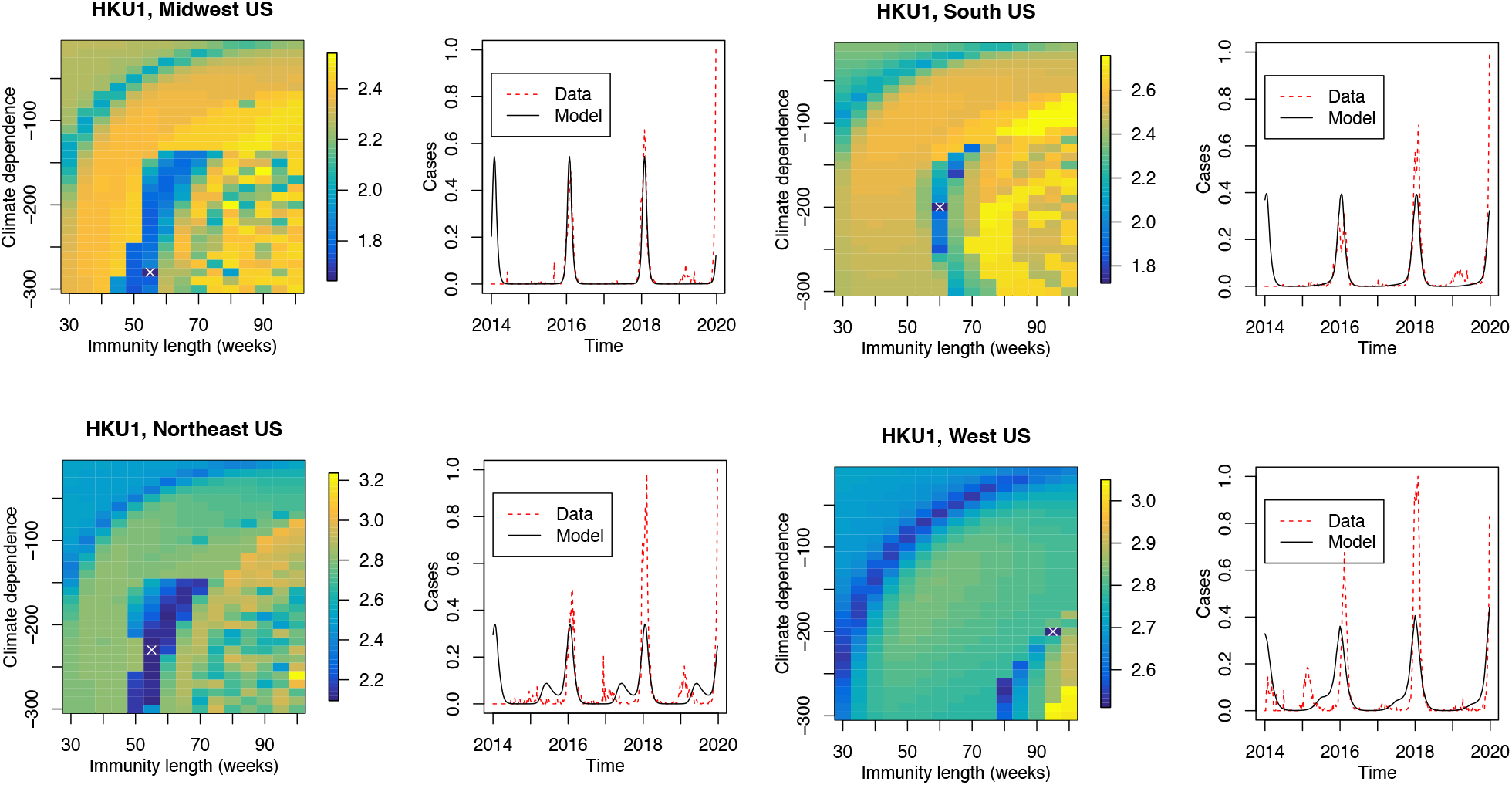
Model fit for HKU1. Two parameters are fitted (climate dependence and immunity length) using case data for HKU1 in four US census regions. Our model fits only the average seasonality of HKU1 cases and does not take into account year-to-year variation. The surface plot shows the log of the sum of absolute error. Darker regions represent a better fit of our model and the best fit parameters are shown with a white cross.

**Figure 3:**
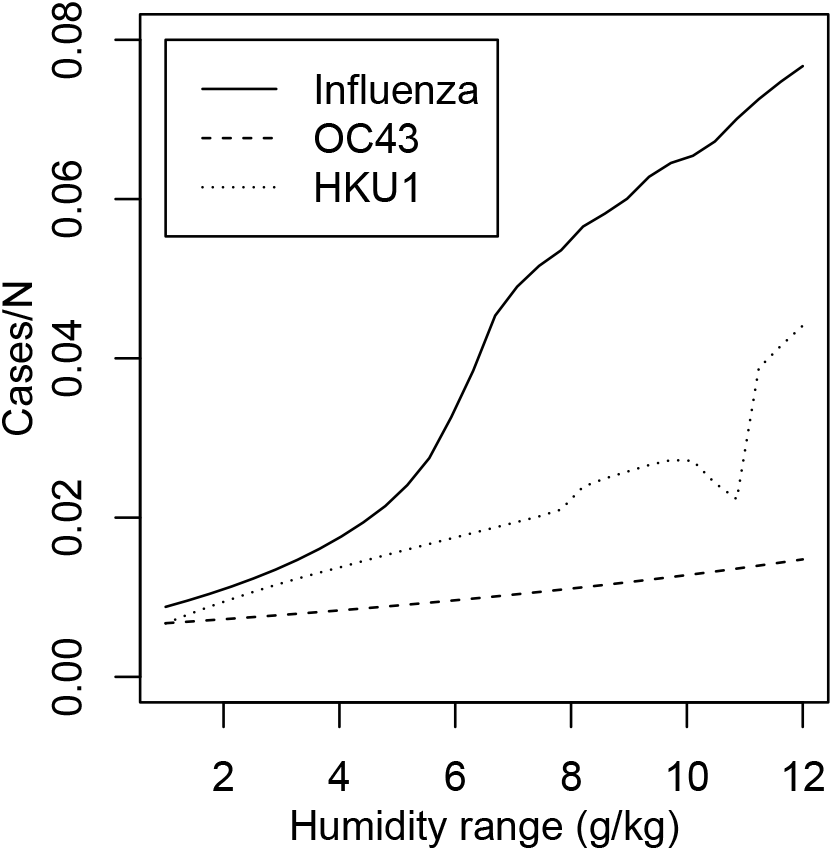
Maximum annual incidence and specific humidity for endemic SARS-CoV-2. The effect of climate (seasonal humidity range) on incidence once the disease has reached stable endemic dynamics.

